# Synthetic Health Data Can Augment Community Research Efforts to Better Inform the Public During Emerging Pandemics

**DOI:** 10.1101/2023.12.11.23298687

**Authors:** Anish Prasanna, Bocheng Jing, George Plopper, Kristina Krasnov Miller, Jaleal Sanjak, Alice Feng, Sarah Prezek, Eshaw Vidyaprakash, Vishal Thovarai, Ezekiel J. Maier, Avik Bhattacharya, Lama Naaman, Holly Stephens, Sean Watford, W. John Boscardin, Elaine Johanson, Amanda Lienau

## Abstract

The COVID-19 pandemic had disproportionate effects on the Veteran population due to the increased prevalence of medical and environmental risk factors. Synthetic electronic health record (EHR) data can help meet the acute need for Veteran population-specific predictive modeling efforts by avoiding the strict barriers to access, currently present within Veteran Health Administration (VHA) datasets. The U.S. Food and Drug Administration (FDA) and the VHA launched the precisionFDA COVID-19 Risk Factor Modeling Challenge to develop COVID-19 diagnostic and prognostic models; identify Veteran population-specific risk factors; and test the usefulness of synthetic data as a substitute for real data. The use of synthetic data boosted challenge participation by providing a dataset that was accessible to all competitors. Models trained on synthetic data showed similar but systematically inflated model performance metrics to those trained on real data. The important risk factors identified in the synthetic data largely overlapped with those identified from the real data, and both sets of risk factors were validated in the literature. Tradeoffs exist between synthetic data generation approaches based on whether a real EHR dataset is required as input. Synthetic data generated directly from real EHR input will more closely align with the characteristics of the relevant cohort. This work shows that synthetic EHR data will have practical value to the Veterans’ health research community for the foreseeable future.

## INTRODUCTION

Recent advances in Big Data and Artificial Intelligence (AI) have provided researchers with unprecedented capability to model complexities of real-world EHR data (*1–5*). Yet legitimate concerns about patient privacy and ethical AI have limited the availability of EHR data within the broader AI research community (*6–8*). The use of synthetic data, which partially mimics the statistical properties of real data (*9*, *10*), reduces the restrictions to sharing research results and analysis across multiple organizations (*11*). Consequently, use of synthetic data can increase the scale and efficacy of research while also reducing potential bias by democratizing research.

The precisionFDA platform was developed by the FDA (Office of Digital Transformation [ODT]/Office of Data, Analytics, and Research [ODAR]) to advance precision medicine and inform regulatory science, including approaches such as bioinformatics pipelines for processing omics data produced by next-generation sequencing (NGS) technology. This secure cloud-based platform offers on-demand scalable computation and data storage, access to reference data, and personalized and shared spaces to enable crowdsourcing and collaboration for improving the evaluation and validation of bioinformatics workflows (*12*). Since its public launch on December 15, 2015, precisionFDA has gained over 6,000 community members worldwide representing NGS instrument manufacturers, NGS-based test providers, standards-making bodies, pharmaceutical & biotechnology companies, healthcare providers, academic medical centers, research consortia, and government agencies. PrecisionFDA engages the public via a discussion forum, featured expert blog posts, app-a-thons, and community challenges.

The precisionFDA challenge framework is one of the platform’s most externally engaging features that enables the broader scientific community to compete in biological data challenges with easily accessible resources for submission testing and validation. To date, 37 community challenges and app-a-thons have been completed on precisionFDA, which generated over 800 submissions and led to novel algorithm development, evaluation, and development of best practices (*12–14*).

As the COVID-19 pandemic progressed, real-world evidence accumulated, demonstrating a variety of risk factors for individual risk of disease severity, including age (*15*), obesity (*16*), cardiovascular disease (*17*), diabetes (*18*), and more (*19*). Predictive models developed using EHR can improve our ability to identify patients at high risk and potentially initiate more aggressive treatment early (*20*, *21*). However, Veterans are a special population that, as compared to the general population, has a higher prevalence of underlying risk factors for severe COVID-19 illness (*22*). Veteran population-specific predictive models are needed to both serve as Veteran-specific clinical decision support and to identify potential additional risk factors within this special population. The VHA data is enriched for individuals whose privacy is a matter of national security concern. Additionally, synthetic COVID-19 patients were demonstrated to be an effective surrogate for a variety of public health and clinical tasks (*23*, *24*). Therefore, synthetic data may play a critical role in modeling COVID-19 and developing AI applications in general within the VHA.

To better understand risk and protective factors for COVID-19 among Veterans, FDA and VHA launched the COVID-19 Risk Factor Modeling Phase 1 Challenge to understand this disease’s impact on the Veteran community. Held from June 2, 2020 to July 3, 2020, the Challenge assessed development of machine learning (ML) models and usefulness of synthetic data for addressing the global pandemic. Here we provide an overview of the Challenge’s results and highlight the strengths and weaknesses of different approaches to synthetic data generation.

## METHODS

The Phase 1 Challenge was hosted on the secure, collaborative, cloud-based precisionFDA platform. It called on the community to leverage ML for developing and evaluating computational models to better understand COVID-19-related health outcomes in Veterans, specifically using these five COVID-19 outcomes: disease status, deceased status, ventilation status, days hospitalized, and days in the intensive care unit (ICU). To reduce security barriers and increase community participation, synthetic data was used to anonymously analyze real data. Synthetic health records for 147,451 synthetic patients were generated using the Synthea synthetic patient generator (*25*) and provided to researchers for modeling.

This synthetic data included conditions, encounters, observations, medications, procedures, and patient demographics. To prevent overfitting and provide a reliable estimate of the model’s performance, synthetic data was split with 80% provided to participants for model training and 20% withheld as a test set for model evaluation. Model performance was evaluated separately for each of the five target COVID-19 outcomes. COVID-19 status, alive or deceased status, and ventilation status predictions were evaluated using the Area Under the Receiver Operating Characteristic (AUROC) metric(*26*). Predictions of days hospitalized and days in the ICU were evaluated using Root-Mean-Square Error (RMSE) and concordance index (c-index) (*27*). Each participating submission was summarized and ranked by the sum of total performance across all five modeling tasks.

Considering recent innovative advances in synthetic data generation methodologies, a follow-up VHA COVID-19 Phase 2 Challenge was conducted. In the Phase 2 VHA COVID-19 Challenge, top-performing participants retrained and evaluated their models on two additional health record datasets: 1) a second synthetic dataset generated using the software package Automatic Brewing of Synthetic Electronic Health Record Data (ABSEHRD) (*28*), which implements CorGAN(*29*), a Generative Adversarial Network; and 2) a de-identified real Veteran health record dataset. Model performance on all three datasets was compared through validation and test AUROC for binary health outcomes as well as test RMSE and c-index for continuous health outcomes. Lastly, we compared feature importance, the Scikits light GBM (LGBM) gain feature importance algorithm, across models within each COVID-19 outcome to determine similarities and differences between algorithms, COVID-19 health outcomes, and dataset relationships.

## RESULTS

The VHA Innovation Ecosystem and FDA’s COVID-19 Risk Factor Modeling Challenge sought to develop and evaluate computational models to predict COVID-19-related health outcomes in Veterans and, in Phase 2, assess the value of synthetic data for research purposes. Figure 1 illustrates the workflow for the Challenge. Phase 1 of the Challenge went live on June 2, 2020 and ran until July 3, 2020. Twenty-one teams submitted 34 entries. Phase 2 of the Challenge ran from May 2021 to January 2022, and enlisted the top three Phase 1 participants to assess the value of synthetic data as a reliable resource for researchers outside the U.S. Department of Veterans Affairs (VA) for model prediction purposes.

**Figure 1.**
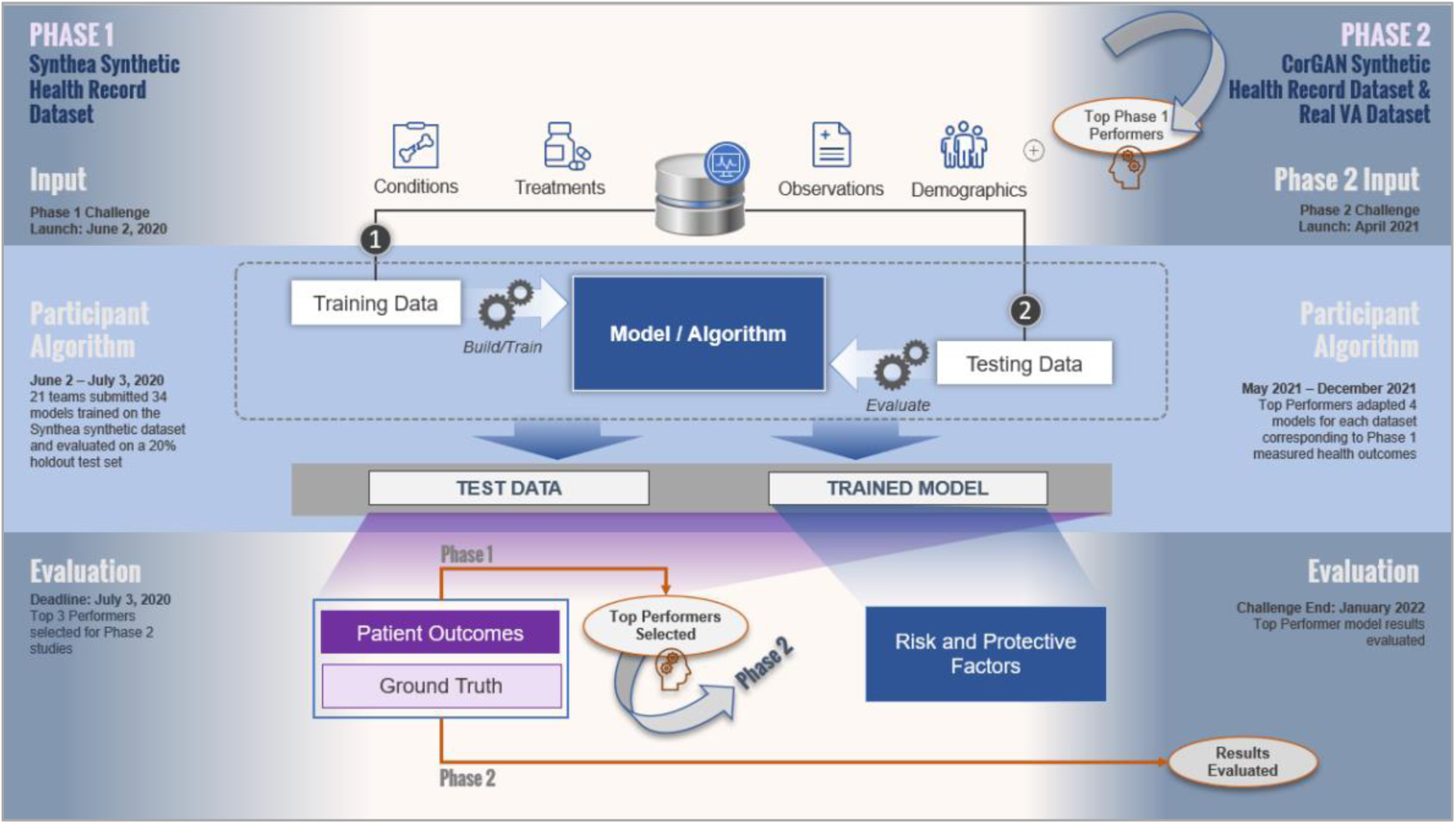
COVID-19 Risk Factor Modeling Challenge Phase 1 Top Performers were Selected to Assist in Synthetic Data Validation in Phase 2. shows the Challenge workflow. In total, 21 teams submitted 34 models trained on the Synthea synthetic dataset and evaluated on a 20% holdout test set. Only the top performing models from Phase 1 were recruited into Phase 2. The focus of Phase 2 was to validate the top performing models on two additional datasets and assess whether synthetic data is a reliable resource for model development to augment community research during emerging pandemics. The CorGAN dataset and Real VA Datasets were added to meet these objectives. Top performing models from Phase 1 were adapted to these datasets using numerous feature engineering processes to remediate differences in naming conventions and units then evaluated on the same metrics as Phase 1. To evaluate models similarly to Phase 1, COVID-19 status was removed from Phase 2 evaluation.

Synthetic data for this challenge were generated through Synthea and CorGAN. The real VA and CorGAN datasets contained 200+ features whereas Synthea included 168 features. Table 1 shows that each of the three datasets used in our analysis had varying levels of accuracy and completeness compared to the Real VA data, including unique distributions across age, sex, and COVID-19 health outcomes. Age was distributed evenly by Synthea reflecting a more realistic generalized population age cross-section, whereas CorGAN distributed age similarly to real VA. Sex was distributed evenly (∼50/50) by Synthea, whereas CorGAN and real VA distributed sex ∼95/5 (male/female). Across COVID-19 binary health outcomes, all datasets distributed similarly with death status being ∼10% of the population and ventilation status being 5%. Across COVID-19 continuous health outcomes, CorGAN and real VA distributed 10% and 20% for ICU and hospitalization, respectively, whereas Synthea had <1% of the population for both health outcomes. An exploratory predictive modeling analysis was performed to guide decisions on input data selection of the Challenge. The exploratory model analysis noted a strong model performance reduction when 2021 data was included in validation thus potentially suggesting a weaker ability to generalize predictions once the COVID-19 vaccine was introduced into the population.

**Table 1.** Synthetic Datasets Demonstrated Varying Efficacy Replicating Age, Sex and Health Outcome Real VA Dataset Distributions. Challenge dataset distributions on age and sex alongside the number of total records, total features, and COVID-19 Challenge Health Outcomes. Synthetic Veteran health records generated by the Synthea Synthetic Population Simulator distributed evenly across age and sex, whereas CorGAN and Real VA Datasets were biased towards male and 60+. Synthea utilizes public knowledge in for the form of health statistics and clinical practice guidelines to parameterize models that generate lifetime synthetic health records. CorGAN synthetic patients were generated using the <u>ABSEHRD package</u>, which utilizes Convolutional Neural Networks to capture the correlations between adjacent medical features in the data representation space by combining Convolutional Generative Adversarial Networks and Convolutional Autoencoders. VA patient records from 2020 in the VA COVID Repository were used as input to generate these results. The Real VA Dataset included ∼72,000 Veterans from 2020 in the VA COVID Repository database with medical history data aligning to the Synthea-generated dataset. This dataset was primarily used as a benchmark to compare the efficacy of alternative synthetic datasets.

|  | Synthea | CorGAN | Real VA |
| --- | --- | --- | --- |
| **Number of records** | 75,417 | 50,019 | 72,373 |
| **Number of features** | 168 | 210 | 204 |
| **Age >90** | 0 | 413 | 2,808 |
| **Age 80-89** | 6,528 | 7,663 | 10,138 |
| **Age 70-79** | 13,010 | 25,244 | 33,976 |
| **Age 60-69** | 11,750 | 12,258 | 21,664 |
| **Age 50-59** | 11,699 | 1,961 | 3,773 |
| **Age 40-49** | 12,000 | 3 | 6 |
| **Age 30-39** | 11,005 | 3 | 5 |
| **Age 20-29** | 10,289 | 3 | 3 |
| **Age 10-19** | 0 | 1 | 0 |
| **Age <10** | 0 | 2,200 | 0 |
| **Female** | 37,221 | 2,347 | 3,493 |
| **Male** | 38,196 | 47,672 | 68,880 |
| **COVID status** | 73,697 | 50,019 | 72,373 |
| **Number of deaths** | 5,569 | 5,829 | 7,564 |
| **Ventilator requirement** | 3,961 | 2,339 | 3,164 |
| **Days required in a hospital** | 111 | 10,524 | 15,126 |
| **Days required in an ICU** | 189 | 4,803 | 6,167 |

Teams submitted predictions (normalized confidence scores between 0 and 1) for COVID-19, ventilation, and alive or deceased status. They predicted the number of days for the outcomes of days hospitalized and days in ICU. Phase 1 submissions were evaluated using various metrics such as the AUROC metric, RMSE, and c-index on a 20% holdout test set. Ranking of the performers was determined by summing ranks of the metric for each outcome.

Phase 1 performance is summarized in Figure 2. The models used by the performers for predicting COVID-19 outcomes (such as COVID-19 status, ventilation status, and deceased status) incorporated advanced ML techniques like Gradient Boosting Machines (GBMs), Random Forest, Adaptive Boost (AdaB), LightGBM, and Neural Networks. Ensemble models, which incorporate several individual models to improve performance, were also used by some performers. The top performers all specifically used GBM ML algorithms, which excel in handling complex high-dimensional data and capturing nonlinear relationships, thus outperforming other participants in Phase 1 of the study.

**Figure 2.**
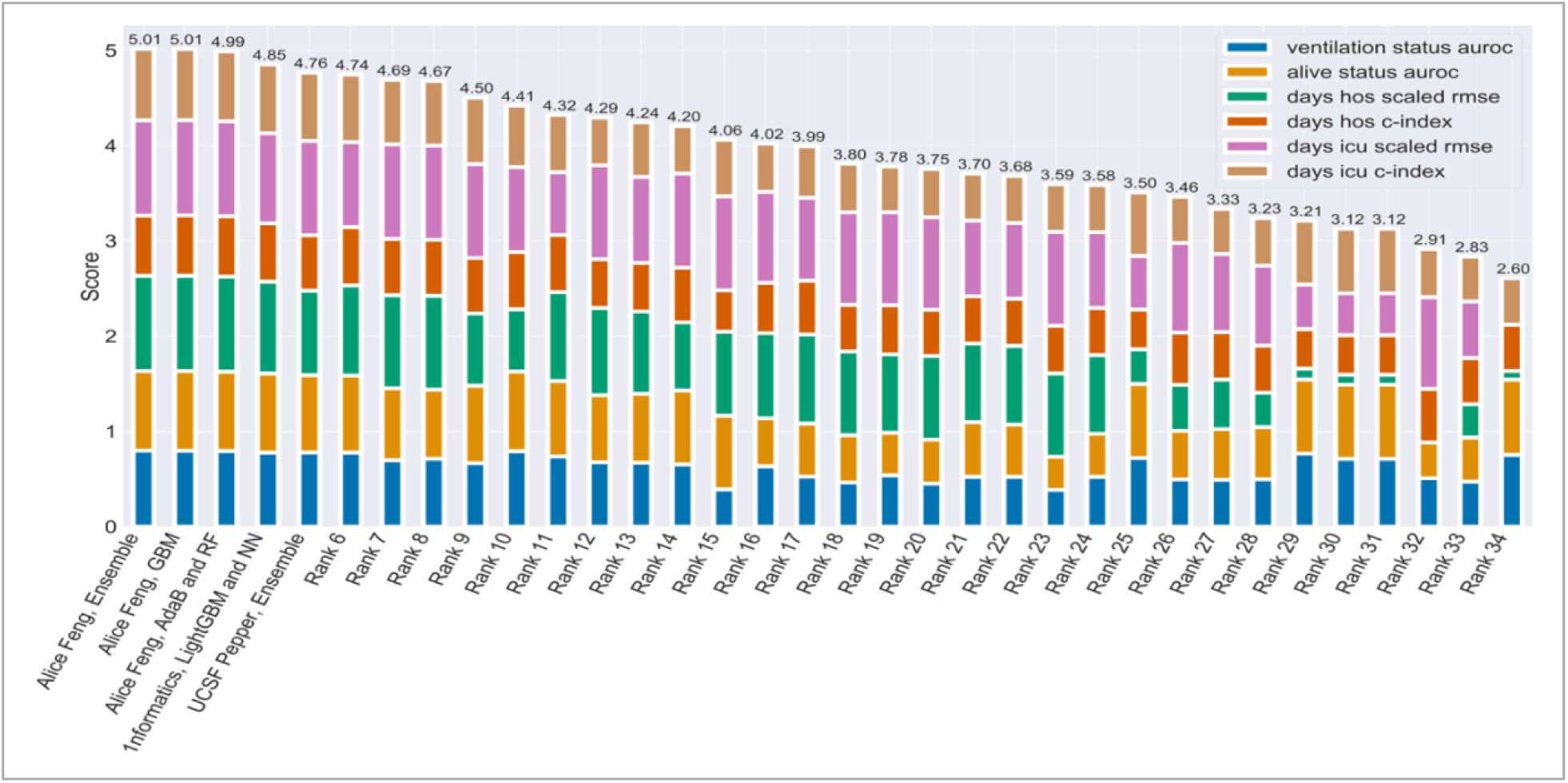
Phase 1 Top Performers Outperformed the Field Through Models Incorporating Gradient Boosted ML. Cumulative bar plot of Phase 1 individual metrics. All submissions except for the top five performers are blinded. The highest score (5.01) represents the best performing model. All metrics except for RMSE are between 0 and 1 where 1 is the best possible score. RMSE was scaled to fit this range using the following equation making 6 as the maximum possible score for Figure 2. All top performing submissions incorporated GBM in the models for binary outcomes, COVID-19 status, ventilation status, and deceased status.

Figure 3 shows that the days hospitalized scaled RMSE metric contained the most variability across all metric and outcome combinations. Interestingly, models that predicted the number of days spent in the ICU exhibited superior RMSE and c-index scores compared to those that predicted days hospitalized with COVID-19 (**Error! Reference source not found.**B and **Error! Reference source not found.**C, respectively).

**Figure 3.**
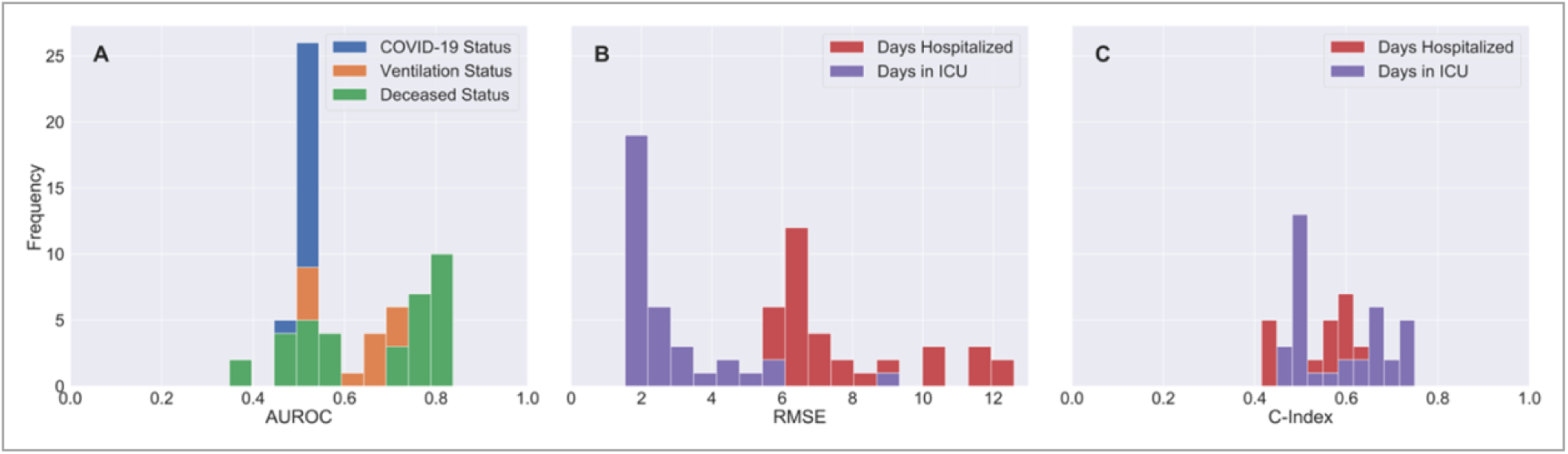
Phase 1 Participant Models Scored Significantly Higher on Severe COVID-19 Health Outcomes on Tracked Metrics. Histograms are displayed for each of the metrics used to evaluate submissions. The average AUROC for COVID-19 status (Figure A) is 0.516 and thus was not considered for identification of top performing models. Models predicting alive or deceased status performed better than models predictive of ventilation status. Models predicting days in ICU had better RMSE and c-indexes than models predicting days hospitalized.

Phase 1 of the Challenge asked the community to develop machine learning models to predict COVID-19-related health outcomes, including COVID-19 status, length of hospitalization, ICU stay, ventilation status, and mortality using synthetic Veteran health records created by the Synthea synthetic patient generator. Across all COVID-19 health outcome model metric distributions, severe COVID-19 health outcomes (such as deceased status and days in ICU) were easier to predict than less severe outcomes (e.g., COVID-19 status, ventilation status, days in hospital). For example, Figure 3A shows that models predicting deceased status had improved performance compared to those predicting ventilation status. Participant model distributions centered >0.50 for ventilation and deceased status but not for COVID-19 status, suggesting that participant models could perform better than random chance at predicting the Synthea dataset outcomes for ventilation and deceased status but not for COVID-19 status. The better performance of models for severe health outcomes (e.g., ventilation status and days in the ICU) may be attributed to the fact that these outcomes have more distinct physiological features (e.g., cardiovascular conditions). In contrast, the COVID-19 status outcome depends on various other factors, such as an individual’s prior medical history, which may not have been a strong predictor of COVID-19 infection status in this dataset.

During Phase 2 of the Challenge, top-performing models from Phase 1 were adapted and validated on additional CorGAN synthetic and real VA datasets. This demonstrated that validation models significantly outperformed random chance in predicting COVID-19 health outcomes during intra-dataset validation. These models were evaluated based on Phase 1 metrics, which included AUROC for binary health outcomes (e.g., alive status and ventilator status), and RMSE for continuous health outcomes (e.g., days in hospital and days in ICU).

The results in Table 2 showed that the Phase 2 top-performer models significantly outperformed random chance predictions on both the synthetic and real Veteran health datasets. The AUROC values for all top participant model predictions on binary outcomes were above 0.5, indicating that the models performed better than random chance in predicting these outcomes. Across all health outcomes, Synthea demonstrated the highest mean metrics of the three datasets in intra-dataset validation. Mean top performer result metrics for the real VA Dataset scored third out of the three datasets in three of the four measured COVID-19 health outcomes in intra-dataset validation. Model result metrics from the Phase 1 winner, Alice Feng, scored the highest of the three teams in seven of the eight metric/outcome combinations.

**Table 2.** Phase 2 Top Performer Validation Models Significantly Outperformed Random Chance. . Phase 2 top performer model validation results in alphabetical order along with mean top performer model results. The top three models from Phase 1 were adapted to each dataset (Synthea, CorGAN and Real VA) and evaluated using Phase 1 metrics on their respective validation datasets. These metrics were AUROC for the binary health outcomes (e.g., alive status and ventilator status), and RMSE for the continuous health outcomes (e.g., days in hospital and days in ICU). Average AUROC across all binary health outcomes were significantly higher than 0.500, suggesting superior model performance against random chance prediction on the validation dataset. Models validated on Synthea data scored the highest across all top performer metrics.

|  | 1Formatics |  |  | Alice Feng |  |  | UCSF Pepper |  |  | Mean |  |  |
| --- | --- | --- | --- | --- | --- | --- | --- | --- | --- | --- | --- | --- |
|  | CorGAN | Synthea | Real VA | CorGAN | Synthea | Real VA | CorGAN | Synthea | Real VA | CorGAN | Synthea | Real VA |
| Alive status (AUROC) | 0.803 | 0.831 | 0.743 | 0.815 | 0.834 | 0.768 | 0.814 | 0.811 | 0.770 | 0.811 | 0.825 | 0.760 |
| Ventilator status (AUROC) | 0.766 | 0.776 | 0.728 | 0.781 | 0.796 | 0.733 | 0.723 | 0.778 | 0.731 | 0.757 | 0.783 | 0.731 |
| Days in Hospitalization (RMSE) | 7.001 | 6.008 | 6.452 | 6.607 | 5.763 | 6.170 | 6.298 | 6.523 | 6.240 | 6.635 | 6.098 | 6.287 |
| Days in ICU (RMSE) | 1.641 | 1.917 | 3.511 | 1.605 | 1.536 | 3.430 | 3.681 | 1.602 | 3.477 | 2.309 | 1.685 | 3.473 |

Top performer model validation results in Phase 2 across all binary health outcomes and dataset combinations demonstrated little variability, suggesting comparable analysis and model development practices among top performers. Figure 4 shows that model validation produced strong binary health outcomes (>0.70 AUROC). Across all health outcomes and datasets, the range of AUROC was <0.06. In five of six health outcomes and dataset pairs, the AUROC range was <0.05; the highest range (0.056) was seen in the ventilator health outcome for CorGAN.

**Figure 4.**
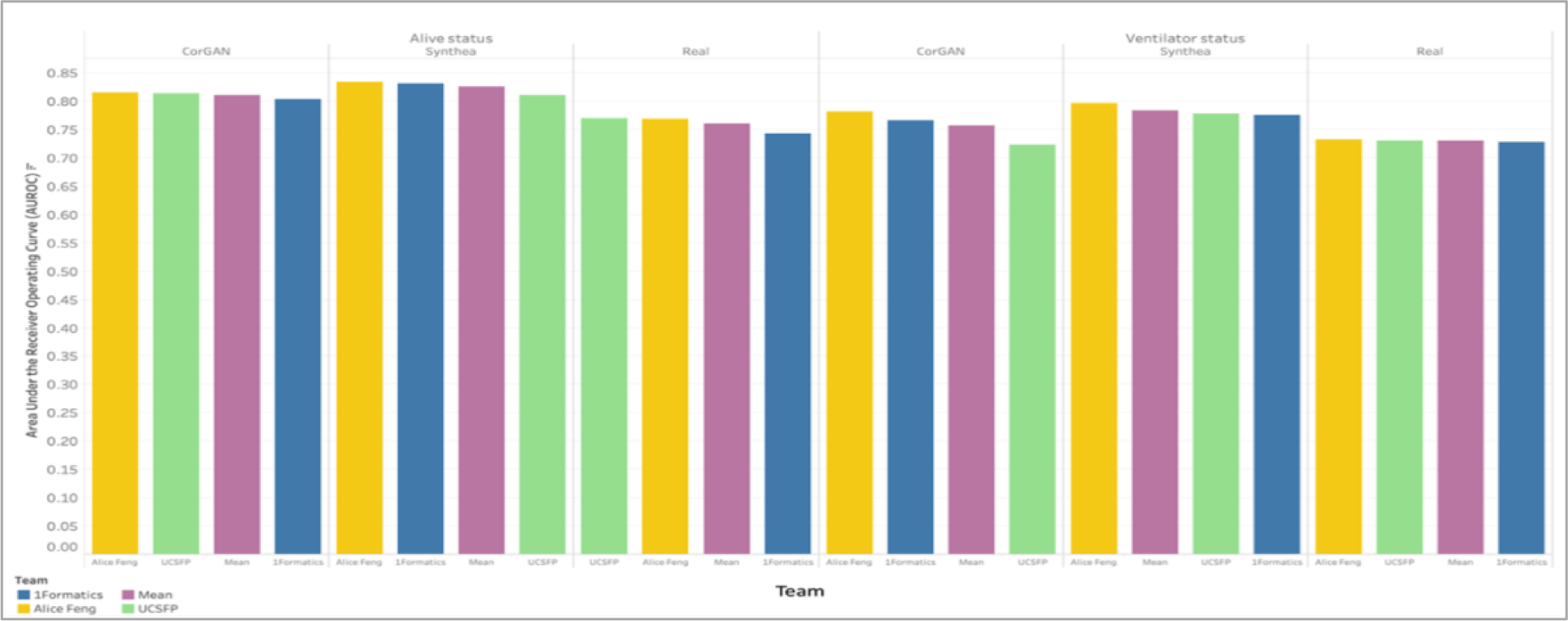
Phase 2 Top Performers Model Demonstrated Minimal Variability Across Outcomes and Datasets. Bar plots are shown for top performer models’ AUROC scores on binary health outcomes (e.g., alive and ventilator status) across datasets (CorGAN, Synthea and Real). Performance was strong on binary health outcomes. Top performers more easily predicted severe outcomes. Real VA Data was the most difficult to predict across both outcomes. Little variability across top performers suggests comparable analysis and model development practices.

All Phase 2 individual models produced significantly superior results on severe COVID-19 health outcomes thus mirroring the pattern seen on models trained and validated on Synthea data. Figure 5 shows that days in ICU ranked Synthea, CorGAN, and real VA data by RMSE performance similarly to the order seen in binary ventilation and deceased status outcomes. Performance was markedly better on severe COVID-19 health outcomes across all models and datasets measured by the RMSE validation metric. Days in hospital outcome ranked Synthea, real VA data, and CorGAN. The range of RMSE for top performers’ models on real data was the smallest of all datasets (0.282 and 0.081).

**Figure 5.**
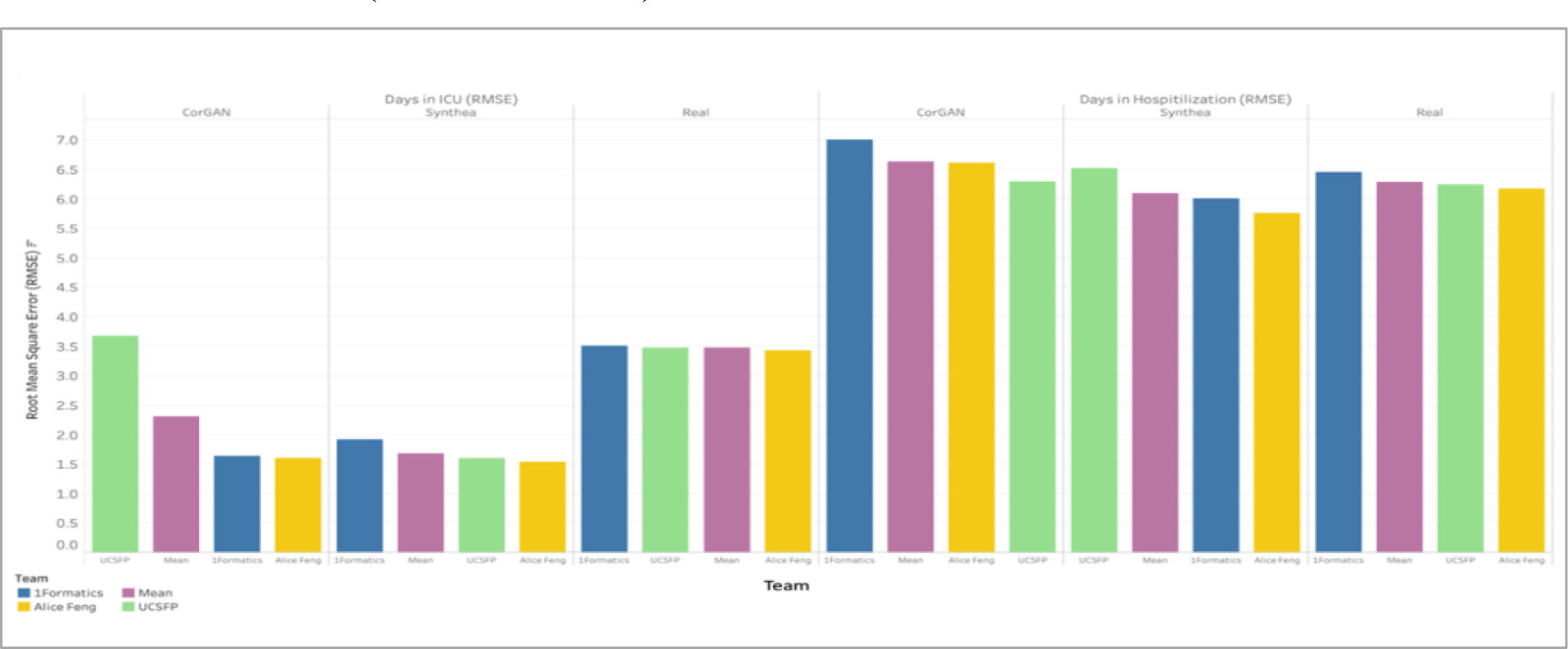
Phase 2 Top Performer Models Performed Significantly Better on Severe COVID-19 Health Outcomes. Bar plots are shown for top performer model performance on COVID-19 continuous health outcomes (e.g., days in ICU and hospitalized) across datasets (CorGAN, Synthea and Real). Top performer models performed better on the more severe outcomes (e.g., days in ICU). Models performed similarly across outcomes and datasets. Similarly, to the binary health outcomes, results were comparable across teams except with days in ICU for the CorGAN dataset.

Risk factor categories marked by a selected Phase 2 top performer varied by dataset and binary health outcome. However, specific risk factor categories were identified as common across datasets and health outcome pairs (i.e., preconditions). All synthetic datasets correctly identified at least one risk factor category that the real VA models also marked. Figure 6 shows that selected top performer models from Phase 2 identified risk factor categories through the Scikits LGBM gain feature importance algorithm. CorGAN models identified risk factor categories that included those identified by models trained and validated on real VA Data (e.g., medication and precondition). CorGAN models identified the most important risk factor category that the real VA models also noted (preconditions) in binary health outcomes (e.g., alive and ventilation status).

**Figure 6.**
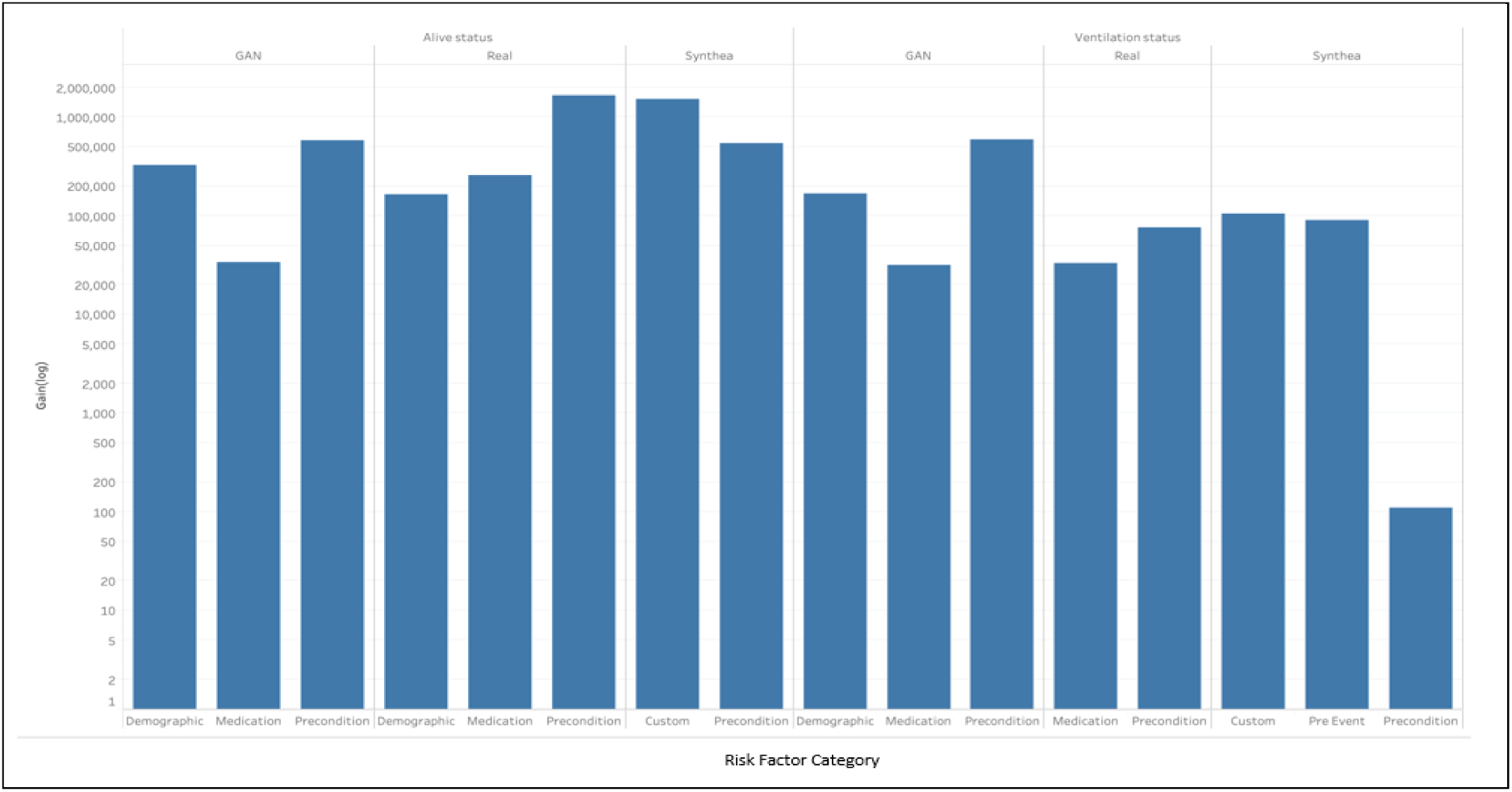
COVID-19 Binary Health Outcome Model Risk Categories Identified in Phase 2 Varied by Dataset. Bar plots are shown for top performers’ model risk factor feature importance by category (e.g., demographic, medication, precondition, pre-event, and custom) across COVID-19 binary health outcomes (death and ventilation status). Categories selected varied by dataset; however, common categories and features were selected across datasets (i.e., preconditions such as diabetes and respiratory failures). Categories selected also varied by health outcome.

Selected top performer models that trained and validated on each synthetic dataset successfully identified at least one of the five most important real VA Data model risk factors including age, septic shock, and dyspnea for each of the binary health outcomes. Table 3 shows that each model identified risk factors commonly linked to COVID-19 (e.g., respiratory, cardiovascular, and chronic health conditions). Features labeled as custom included transformations across multiple features. Risk factors identified by synthetic data were still commonly linked to COVID-19 despite not being identified by the real VA trained and validated models (e.g., kidney conditions and diabetes).

**Table 3.**
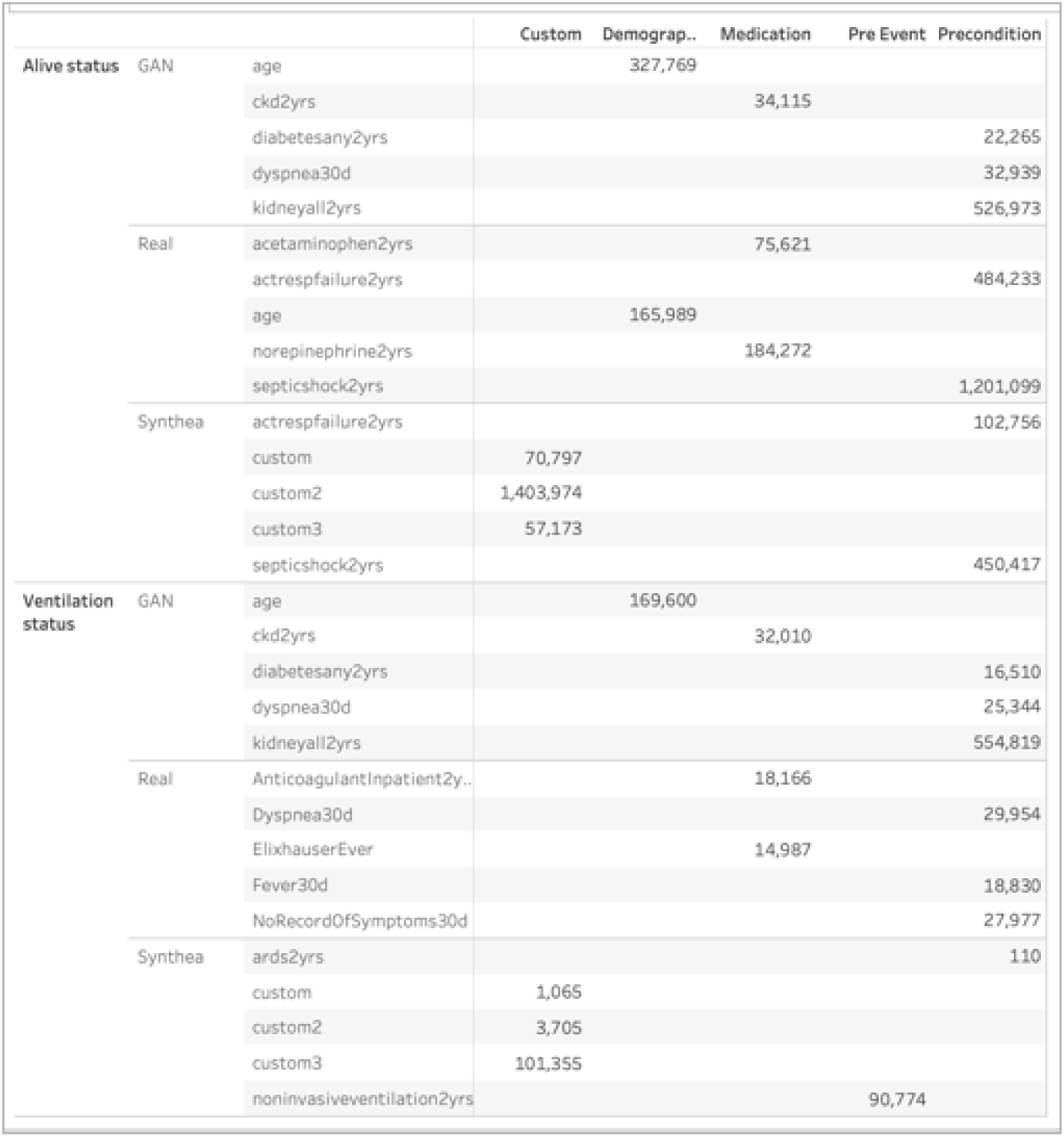
COVID-19 Binary Health Outcomes Identified Common Model Risk Factors Across Datasets in Phase 2. presents Phase 2 top performer risk factor table outlines the top five ranked features for each model by dataset for the binary COVID-19 health outcomes (e.g., alive status and ventilation status). Three out of four synthetic dataset trained models identified at least one risk factor (e.g., age, septic shock, dyspnea) from the model trained on Real data. All models identified risk factors that have been linked to COVID-19 (e.g., respiratory, cardiovascular, and chronic health conditions).

**Table 4.**
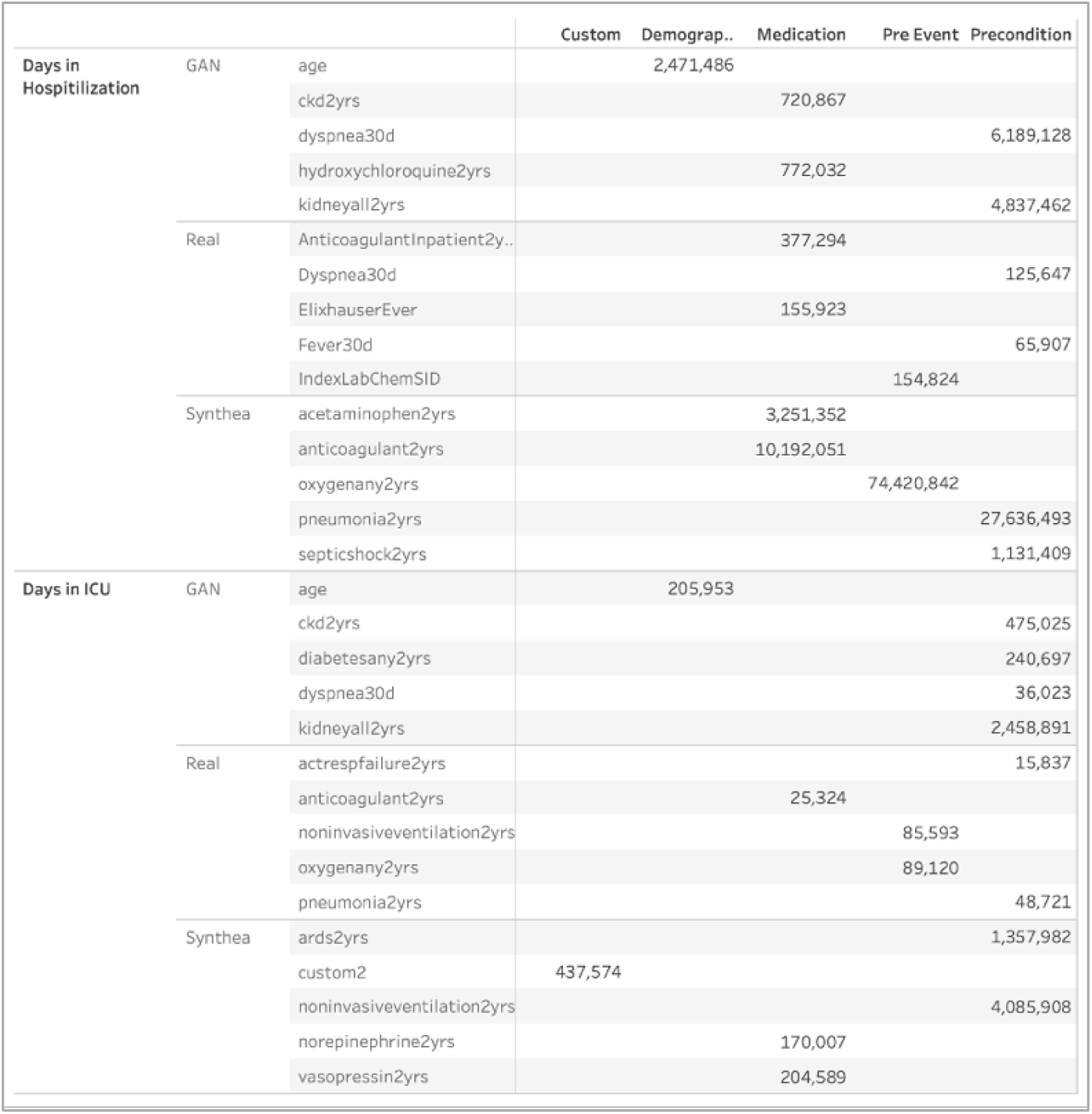
COVID-19 Continuous Health Outcomes Identified Common Model Risk Factors Across Datasets in Phase 2. represents Phase 2 top performer risk factor table outlines the top five ranked features for each model by dataset for the continuous COVID-19 health outcomes (e.g., days in hospital and days in ICU). Three out of four synthetic dataset trained models identified risk factors (e.g., dyspnea, anticoagulant, and non-invasive ventilation) that were also deemed important by models trained on the respective Real dataset. All models identified risk factors that have been linked to COVID-19 (e.g., respiratory, cardiovascular, and chronic health conditions).

Risk Factor categories noted by a selected Phase 2 top performer marked multiple risk factor categories across multiple dataset and health outcome pairs (e.g., medications, preconditions, and pre-events) as shown in Figure 7. For days hospitalized, Synthea correctly identified all three real VA model risk factor categories while CorGAN identified two (medication and precondition). Precondition was correctly identified as a risk factor category across all dataset and continuous health outcome pairs. Each dataset and continuous health outcome pair demonstrated a different gain distribution.

**Figure 7.**
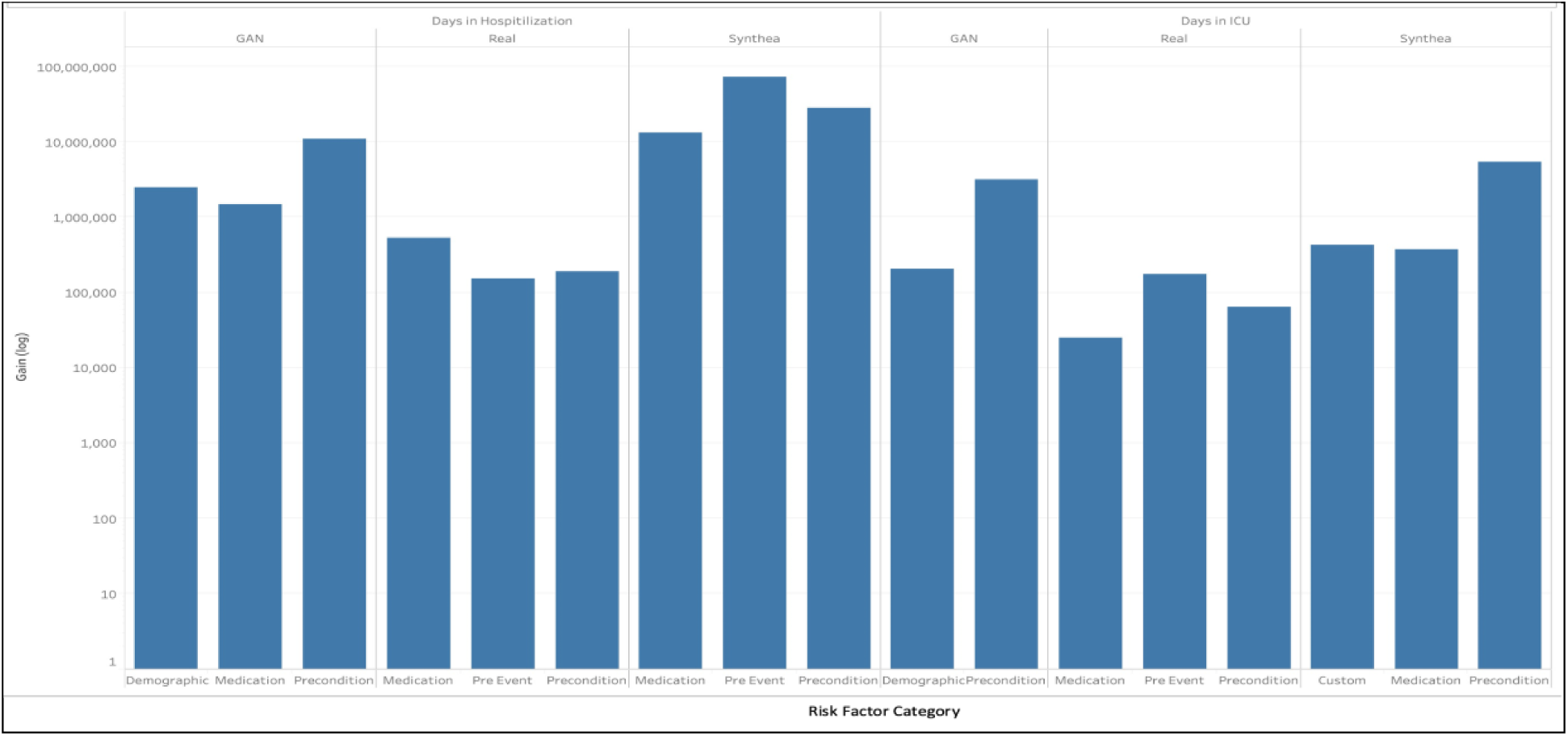
COVID-19 Continuous Health Outcome Models Identified Common Risk Categories in Phase 2. Bar plots are shown for a top performers’ model risk factor feature importance by category (e.g., demographic, medication, precondition, pre-event, and custom) across COVID-19 continuous health outcomes (e.g., days in hospital and days in ICU). Risk factor categories selected varied by dataset; however, common categories and features were selected across datasets. All models trained on synthetic datasets correctly identified the preconditions risk factor category and three out of four synthetic datasets identified precondition and medication noted by the model trained on Real data. Categories selected varied by health outcome.

Selected top performer models trained and validated on each synthetic dataset successfully identified at least one of the five most important real VA data model risk factors including non-invasive ventilation, dyspnea, and anticoagulant for each continuous health outcome. Error! **Reference source not found.. COVID-19 Continuous Health Outcomes Identified Common Model Risk Factors Across Datasets in Phase 2** represents shows that each model identified risk factors commonly linked to COVID-19 (e.g., respiratory, cardiovascular, and chronic diseases as well as general health conditions). Risk factors identified by synthetic data were still commonly linked to COVID-19 despite not being identified by the real VA trained and validated models (e.g., kidney conditions and diabetes).

## DISCUSSION

Our study demonstrated that ML models for predicting COVID-19 health outcomes in Veteran populations trained on synthetic data have similar properties to those trained on real data. For CorGAN data generated with ABSEHRD, we noted similarities in both demographic and COVID-19 health outcomes cross tabulations with real data. In contrast to CorGAN, the Synthea data was generated based on a different cohort and, therefore, did not align with demographic variables yet still matched the COVID-19 health outcome distributions. Modeling results were similar across the three datasets including top performer rankings, quantitative performance metrics, and feature importance metrics. Important features identified by each model were known COVID-19 severity risk factors. We conclude that the underlying similarity between real and synthetic data generated by packages such as ABSEHRD and Synthea have real practical value for research on Veterans’ health.

However, clear differences existed between the datasets and further research is needed to quantify the degree of efficacy in fully replicating real data. Our results demonstrate that models trained on synthetic data show systematically inflated performance metrics compared to those trained on real data. The inflation of performance metrics is indicative of a loss of data complexity and variability during the synthetic generation process—in other words, modeling real data is harder than modeling synthetic data. Further improvements in synthetic data-generating algorithms and available training datasets may help with shrinking this gap in the future. A limitation of our study design is that we did not test the synthetic data trained models on real datasets. Future studies should be designed to directly test whether synthetic data can substitute for real data in this way. Another limitation of this study is that the variety of ML algorithms used was low. Because of the challenge format, the top performers happened to all select Gradient Boosted Machines and while this may be indicative of which models work well for this data, it prevented an assessment of whether all algorithms are affected similarly by the differences between real and synthetic EHR data.

Fundamental tradeoffs exist when selecting an approach for crowdsourcing ML model development with EHR data. Although real EHR data is the most austere, the barrier to access will limit researcher participation. Packages like ABSEHRD are publicly available yet still require access to a real patient data cohort to generate synthetic data. The benefit of using real patient data from the cohort of interest is that synthetic data can more closely match the real data. Synthea does not require access to a real patient dataset because the developers have pre-populated population modules spanning numerous segments of the U.S. population. Therefore, anyone can generate synthetic EHR data with Synthea although it may not represent the real cohort of interest or data directly generated from that cohort. The pros and cons of each approach should be weighed on a case-by-case basis.

In conclusion, the Phase 2 Challenge promoted development and documentation of applying cutting-edge ML methodologies to better understand the emerging COVID-19 pandemic. The challenge results provide a public resource for comparing synthetic data generation methodologies and the efficacy of ML algorithms against those respective techniques. Continued improvements in synthetic data-generating algorithms and training datasets will shrink the gap between real and synthetically trained ML models.

## ETHICS STATEMENT

The San Francisco General Hospital (SFGH) Committee Institutional Review Board (IRB), affiliated with SFGH and University of California San Francisco (UCSF) reviewed and waived the need for ethics approval.

## Data Availability

All data produced are available online at precision.fda.gov

https://precision.fda.gov/challenges/11

## ACKNOWLEDGEMENTS

Bocheng Jing and John Boscardin were supported by NIH grants P01AG066605 and P30AG044281.

